# Machine Learning-Based Prediction of Coronary Care Unit Readmission: A Multi-Hospital Validation Study

**DOI:** 10.1101/2024.03.19.24304553

**Authors:** Fei-Fei Flora Yau, I-Min Chiu, Kuan-Han Wu, Chi-Yung Cheng, Wei-Chieh Lee, Huang-Chung Chen, Cheng-I Cheng, Tien-Yu Chen

**Author notes:** Corresponding author: Tien-Yu Chen, address: No. 123, Dapi Rd., Niaosong Dist., Kaohsiung City, Taiwan, 83301.

## Abstract

Readmission to the Coronary Care Unit (CCU) has significant implications for patient outcomes and healthcare expenditure, emphasizing the urgency to accurately identify patients at high readmission risk. This study aims to construct and externally validate a predictive model for CCU readmission using machine learning (ML) algorithms across multiple hospitals. Patient information, including demographics, medical history, and laboratory test results were collected from electronic health record system and contributed to a total of 40 features. Three ML models, Logistic Regression, Random Forest, and Gradient Boosting were employed to estimate the readmission risk. The gradient boosting model was selected demonstrated superior performance with an Area Under the Receiver Operating Characteristic Curve (AUC) of 0.887 in the internal validation set. Further external validation in hold-out test set and three other medical centers upheld the model’s robustness with consistent high AUCs, ranging from 0.852 to 0.879. The results endorse the integration of ML algorithms in healthcare to enhance patient risk stratification, potentially optimizing clinical interventions and diminishing the burden of CCU readmissions.

**Key learning points:** What is already known:

1. Readmission to the CCU has significant implications for both patient outcomes and healthcare costs.
2. Accurately distinguishing patients at high or low risk for CCU readmission is essential for clinicians to allocate resources effectively

What this study adds:

1. A predictive model for CCU readmission was constructed using machine learning algorithms trained from one medical center and validated externally in three major medical centers.
2. Among the ML models evaluated, the Gradient Boosting model showed the highest performance with an AUC of 0.879 in hold-out test set, and its robustness was further confirmed in external validation across three medical centers with an AUC range from 0.848-0.863.
3. By using different cut-off thresholds to prioritize the model’s sensitivity or specificity, clinicians can distinguish between high-risk and low-risk patients, enabling them to determine the appropriate level of monitoring and treatment planning for those at high risk.

## Introduction

Readmission to coronary care unit (CCU) after initial hospitalization is a critical problem that has significant implications for patient outcomes and healthcare costs. Patients who are readmitted to intensive care unit have worse outcomes, including higher mortality rates, longer hospital stays, and increased healthcare costs^1^. According to previous studies, the readmission rate to CCU ranges from 2.5% to 8% before discharge of initial hospitalization^2–4^.

Identifying patients at high risk of readmission is essential for improving patient outcomes and reducing healthcare costs. By identifying high-risk patients, healthcare providers can implement targeted interventions, such as medication adjustments, lifestyle modifications, and care coordination, to prevent readmission and improve outcomes. Therefore, developing accurate predictive models for readmission risk is critical for improving patient care and reducing healthcare costs.

Machine learning (ML) has emerged as a powerful tool in healthcare for predicting various outcomes and improving patient care. ML algorithms can detect complex patterns and relationships in data that may not be apparent to humans, making it a valuable tool for predicting readmission after discharge from CCU. Previous studies have used ML to predict readmission risk in various patient populations, including those with heart failure, chronic obstructive pulmonary disease, and diabetes^5–7^ .

The objective of this study is to develop a predictive model for readmission to CCU after initial hospitalization using ML algorithms. Specifically, we aim to identify patient characteristics and clinical factors that are associated with readmission and develop a model that can accurately predict readmission risk.

## Method

### Study setting

In this prognostic study, we leveraged electronic health records from Lin Kou Chang Gung Memorial Hospital (derivation cohort) collected from October 1, 2000, to June 30, 2019, to establish the development cohort. For external validation, we included records from three major medical centers located diversely nationwide. These centers are Kaohsiung Chang Gung Hospital (validation cohort A), Chiayi Chang Gung Hospital (validation cohort B), and Keelung Chang Gung Hospital (validation cohort C). The data spanned from January 1, 2001, to June 30, 2019. All four hospitals are recognized as the largest healthcare facilities in their respective local areas, ensuring a diverse and comprehensive dataset for this study. Prior to the commencement of the study, ethical approval was obtained from the Institutional Review Board of Chang Gung Medical Foundation. The approval reference number is 202201541B0.

### Data collection

In the development cohort, we included all patients admitted to the CCU during the study period. We excluded patients who died or were transferred to other hospitals during their CCU admission. The complete dataset was randomly partitioned into three subsets at an 8:1:1 ratio, designated as the training, internal validation, and hold-out test sets, respectively. To ensure the integrity of the data, we used patient-level splitting; this approach guaranteed that different admissions of the same patient were not included in multiple subsets (Figure 1).

For this study, we collected a total of 40 features from the Electronic Health Records (EHR), encompassing patient demographic characteristics, laboratory test results, vital signs, and past medical history, all of which are relevant to the prognosis during the CCU admission. Diagnostic information upon and during CCU admission, as well as cardiovascular treatments, were also gathered as predictive features (see Table 1). The primary outcome was CCU readmission, defined as a return to the CCU or death occurring post-discharge from the CCU within the same hospital stay.

**Figure 1.**
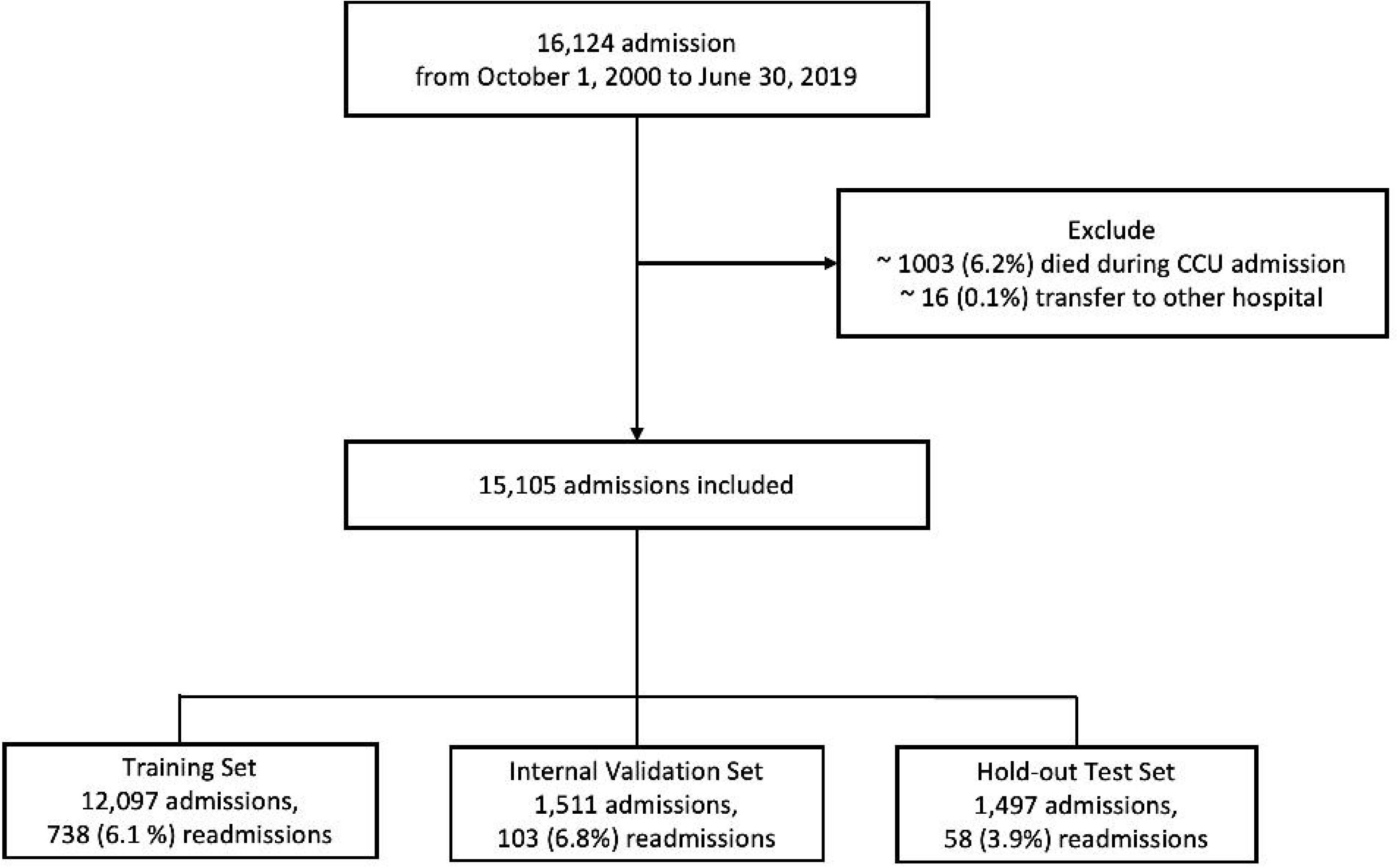
Patient inclusion flowchart of this study.

**Table 1.** Patient’s demographics of included Hospitals.

| Variable | Derivation Cohort<br>n=15105<br>N(%) / Mean(SD) | Validation Cohort A<br>n=9651<br>N(%) / Mean(SD) | Validation Cohort B<br>n=8821<br>N(%) / Mean(SD) | Validation Cohort C<br>n=6608<br>N(%) / Mean(SD) |
| --- | --- | --- | --- | --- |
| Sex |  |  |  |  |
| male | 9796(64.9%) | 6715(69.6%) | 5378(61%) | 4024(60.9%) |
| female | 5309(35.1%) | 2936(30.4%) | 3443(39%) | 2584(39.1%) |
| Age | 66.2(14.6) | 65.9(13.5) | 70.7(14) | 69.4(14.6) |
| Main Diagnosis |  |  |  |  |
| ACS | 9158(60.6%) | 6640(68.8%) | 4466(50.6%) | 3385(51.2%) |
| OHCA | 77(0.5%) | 50(0.5%) | 156(1.7%) | 23(0.3%) |
| IHCA | 643(4.2%) | 567(5.9%) | 465(5.3%) | 381(5.8%) |
| Readmission | 899(6%) | 520(5.4%) | 569(6.5%) | 409(6.2%) |

ACS=Acute Coronary Syndrome; OHCA=Out-of-Hospital Cardiac Arrest; IHCA=In-Hospital Cardiac Arrest;

For the laboratory tests, we gathered the standard tests conducted during the same admission and selected the test closest to the CCU discharge for analysis. For the left ventricular ejection fraction (LVEF), obtained from echocardiography reports within one year, we used the measurement nearest to the CCU discharge. Information on procedures and events was collected and identified in relation to the same episode of CCU admission.

Missing values were observed in the numerical features, with missing rates ranging between 1.2% and 4.5%. These missing values were imputed using the median value from the remaining data. Before model training, all feature values were normalized to a scale of [0,1], taking into account the categorical features.

### Development of Machine Learning Models

In this study, we employed three machine learning models, namely Logistic Regression (LR), Random Forest (RF), and Gradient Boosting (GB) to predict the risk of patients being readmitted to the CCU. LR is a fundamental statistical model widely employed in machine learning for binary classification tasks^8–10^. By examining a cohort’s clinical predictors, the LR model estimates the probability of readmission by assigning weights to input features. The model’s output is a value between 0 and 1, interpreted as the likelihood of an event occurring, in this study, the return of a patient to CCU. This method is particularly valued for its simplicity, making it a baseline standard tool for predictive analytics. RF is a type of ensemble learning algorithm that is based on the decision tree method. It employs bagging, a sampling technique that randomly selects subsets of the training data and features to create multiple classification and regression trees. The results of the individual trees are then aggregated through a voting process to arrive at the final prediction. The idea behind this technique is that each tree is slightly different due to the random selection of data and features, which reduces the risk of overfitting to the training data. In various medical studies, random forest has shown higher predictive power that has been widely adopted for predicting risk outcomes and predictive factors^11–13^. GB operates by sequentially building an ensemble of weak learners, each one focusing on correcting the errors of its predecessor. Each weak learner, typically a decision tree, is added to the ensemble with the goal of reducing the loss function, a measure of how well the model fits the entire dataset^14^. The performance of each tree in the sequence is taken into account by optimizing an objective function that combines the predictions with the actual values. This technique often results in a powerful predictive model that can handle various types of data and different distributions. Gradient Boosting has been widely recognized for its effectiveness in classification and regression tasks, often achieving high accuracy and robustness in predictive analytics challenges.

We incorporate feature selection process during training ML model in this study. We adopted a forward stepwise feature selection algorithm that help to select the optimal subset of features for each ML models. Forward stepwise feature selection is a process used in model building that begins with an empty model and adds in features one by one. In each step, the feature that provides the most significant improvement to the model fit is included, until the addition of new feature no longer significantly improves model performance^15^. The rationale behind choosing the forward stepwise feature selection method is multifactorial. First, it allows for a deliberate escalation in model complexity and ensures that each feature incorporated plays a pivotal role in the predictive accuracy of the model. Second, it prevents overfitting thereby bolstering the model’s generalizability, and it augments interpretability by focusing only on the most important features. This feature selection method is particularly useful when dealing with datasets with a large number of variables.

### Statistical Analysis

Continuous variables are expressed as mean ± standard deviation, while nominal variables are presented as proportions. Ordinal variables are depicted as median along with their interquartile range (IQR) or mean (standard deviation) if they were normally distributed. The comparative performance of three machine learning algorithms, LR, RF, and GB was assessed with the Area Under the receiver operating characteristic Curve (AUC) and F-1 score serving as the primary evaluation metric. Upon selecting the optimal model through hold-out test set, its robustness was further evaluated in different medical centers, with performance metrics including sensitivity, specificity, positive predictive value (PPV), and negative predictive value (NPV). All analyses were conducted using Python 3.8, utilizing the Scikit-Learn package.

## Results

The demographic data for all included hospitals are presented in Table 1. Our model was trained using data from derivation cohort, which had the highest patient count of 15,105, followed by validation cohort A with 9,651 patients, validation cohort B with 8,821, and validation cohort C with 6,608. In derivation cohort, there was a male predominance of 64.9% (9,796 patients), and the mean age was 66.2±14.6 years. The primary diagnosis was acute coronary syndrome (ACS), accounting for 60.6% of cases, and the readmission rate was 6%. For the three medical centers used as external validation, the proportion of male patients ranged from 60.9% to 69.6%, the mean patient age varied from 65.9 to 70.7 years, and the readmission rates across these hospitals varied from 5.4% to 6.5%. The complete list of variables collected is shown in Supplemental table 1.

The outcome of the forward stepwise feature selection algorithm for determining the optimal subset of predictive features for CCU readmission is delineated in Table 2. Across the three machine learning models analyzed, the use of a ventilator and the length of CCU stay were consistently selected as the first and second most significant features, respectively. For the LR model, platelet count, chronic obstructive pulmonary disease (COPD), sodium levels, intra-aortic balloon pump (IABP) use, in-hospital cardiac arrest (IHCA), and creatinine levels were identified as the subsequent predictors in order of importance. The GB model included additional predictors such as hypertension, peripheral artery occlusive disease (PAOD), myocardial infarction (MI), extracorporeal membrane oxygenation (ECMO) usage, coronary artery disease (CAD), diabetes mellitus (DM), and atrial fibrillation. The RF model identified COPD, ECMO, and out-of-hospital cardiac arrest (OHCA) as other important features. Notably, certain predictors like IABP and COPD were selected by multiple models across different modeling approaches.

**Table 2.** Result variables of Stepwise features selection.

|  | Logistic Regression | Gradient Boosting | Random Forest |
| --- | --- | --- | --- |
| 1<sup>st</sup> feature | Ventilator | Ventilator | Ventilator |
| 2<sup>nd</sup> feature | Length of CCU stay | Length of CCU stay | Length of CCU stay |
| 3<sup>rd</sup> feature | Platelet | Hypertension | COPD |
| 4<sup>th</sup> feature | COPD | PAOD | ECMO |
| 5<sup>th</sup> feature | Sodium | MI | OHCA |
| 6<sup>th</sup> feature | IABP | IABP |  |
| 7<sup>th</sup> feature | IHCA | COPD |  |
| 8<sup>th</sup> feature | Creatinine | ECMO |  |
| 9<sup>th</sup> feature |  | CAD |  |
| 10<sup>th</sup> feature |  | DM |  |
| 11<sup>th</sup> feature |  | Atrial Fibrillation |  |
| 12<sup>th</sup> feature |  | OHCA |  |

COPD=Chronic Obstructive Pulmonary Disease; PAOD=Peripheral Arterial Occlusive Disease; ECMO=ExtraCorporeal Membrane Oxygenation; MI=Myocardial Infarction; OHCA=Out-of-Hospital Cardiac Arrest; IABP= Intra-aortic balloon pump; IHCA=In-Hospital Cardiac Arrest; CAD=Coronary Artery Disease; DM=Diabetes Mellitus

The performance of the three models, as per their respective AUCs, was commendable with scores of 0.858, 0.875, and 0.887 in the internal validation set, for LR, RF, and GB, respectively (Figure 2A). Correspondingly, their F-1 scores were 0.253 for LR, 0.271 for RF, and 0.284 for GB. Accounting for performance, the GB model was chosen to predict the risk of CCU readmission in hold-out test set and external validation. In this study, external validations were conducted on patient data procured from diverse geographical regions. The model exhibited significant discriminatory ability in predicting CCU readmissions, supported by consistent high AUCs in test set of 0.879, and from validation cohort A, B, and C, with values of 0.863, 0.848, and 0.852, respectively, as illustrated in Figure 2B.

**Figure 2.**
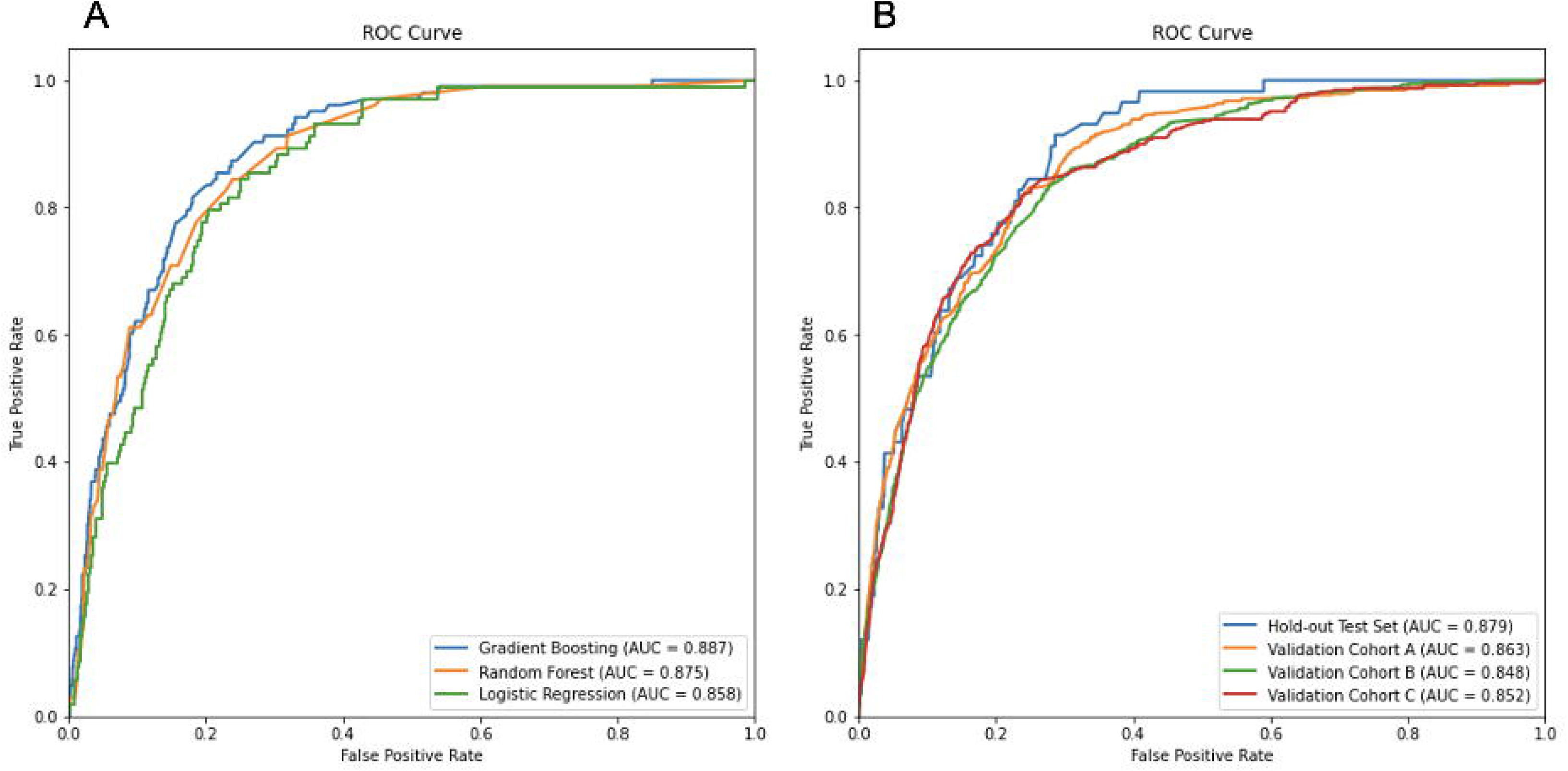
ROC curve and AUC value of different machine learning models in internal validation (2A). ROC curve and AUC value of Gradient Boosting in hold-out test set and external validation (2B).

For binary classification, the default decision threshold is set to 0.5. However, this could be adjusted to substantially impact the model’s performance owing to the trade-off between false positives and false negatives. Table 3 demonstrates the model’s performance in predicting CCU readmission at different cutoff values within the hold-out test set and external validation sets from validation cohort A, B, and C. The threshold of 0.025, identified by the Youden Index as optimal, yields a balance with a sensitivity of 0.915 and specificity of 0.724 in the hold-out test set, accompanied by a PPV of 0.154 and NPV of 0.995. The selection of cutoff values at 0.125 is strategic, aiming to attain a specificity of 0.9 in the hold-out test set. At a cutoff value of 0.125, specificity is prioritized, while sensitivity decreases to 0.534, and PPV rises to 0.233, and NPV of 0.969. The respective sensitivity, specificity, PPV, and NPV to three external validation hospital were demonstrated in Table 3.

**Table 3.**
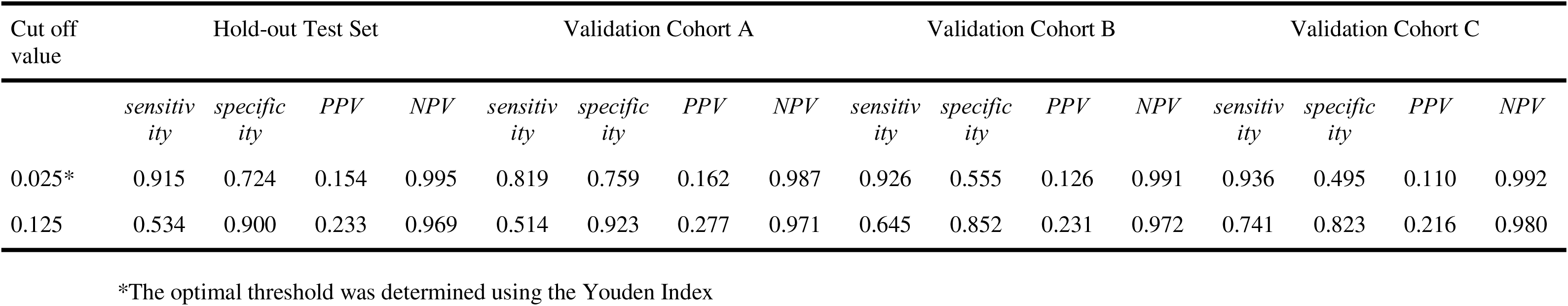
Model performance on predicting CCU readmission in different cut-off value.

| Cut off value | Hold-out Test Set |  |  |  | Validation Cohort A |  |  |  | Validation Cohort B |  |  |  | Validation Cohort C |  |  |  |
| --- | --- | --- | --- | --- | --- | --- | --- | --- | --- | --- | --- | --- | --- | --- | --- | --- |
|  | *sensitivity* | *specificity* | *PPV* | *NPV* | *sensitivity* | *specificity* | *PPV* | *NPV* | *sensitivity* | *specificity* | *PPV* | *NPV* | *sensitivity* | *specificity* | *PPV* | *NPV* |
| 0.025* | 0.915 | 0.724 | 0.154 | 0.995 | 0.819 | 0.759 | 0.162 | 0.987 | 0.926 | 0.555 | 0.126 | 0.991 | 0.936 | 0.495 | 0.110 | 0.992 |
| 0.125 | 0.534 | 0.900 | 0.233 | 0.969 | 0.514 | 0.923 | 0.277 | 0.971 | 0.645 | 0.852 | 0.231 | 0.972 | 0.741 | 0.823 | 0.216 | 0.980 |

\*The optimal threshold was determined using the Youden Index

## Discussion

The development of a machine learning model for predicting the risk of readmission to the Coronary Care Unit (CCU) before discharge from the initial hospitalization represents a significant advancement in healthcare practice. The study’s findings suggest that when ML algorithms are properly trained and validated, they can provide an accurate and reliable method for identifying patients at risk of readmission. Machine learning is particularly adept at analyzing complex patterns and relationships in data, which makes it an ideal tool for predicting the risk of readmission. In this study, three machine learning models—Logistic Regression (LR), Random Forest (RF), and Gradient Boosting (GB)—performed exceptionally well, with AUCs ranging from 0.858 to 0.887. These results underscore the potential of machine learning in healthcare, especially in predicting readmission risk and identifying high-risk patients. Furthermore, we established two different thresholds in this study, which prioritize either high sensitivity or high specificity. By utilizing these thresholds, healthcare providers can implement targeted interventions, such as continuous monitoring in a general ward or intensive medical treatment, to mitigate the risk of CCU readmission, thereby improving patient outcomes and reducing healthcare costs.

ML algorithms can use a variety of data sources to predict readmission risk, including electronic health records, clinical notes, and demographic data. ML algorithms can also incorporate non-traditional data sources, such as social determinants of health, to improve the accuracy of predictions^16^. By incorporating multiple data sources, ML algorithms can develop more accurate predictive models and identify high-risk patients who may benefit from targeted interventions to prevent readmission. However, there are also challenges associated with using ML in healthcare. One major challenge is the need for high-quality data. ML algorithms require large amounts of data to develop accurate predictive models, and the quality of the data is critical for ensuring that the models are reliable and valid. In addition, there are concerns about bias in ML algorithms, as they may perpetuate existing disparities in healthcare if they are trained on biased data^17^.

Predicting the trajectory of patients recovering after leaving CCU is challenging. A two-year study found that a quarter of patients experienced mortality after readmission to the CCU^18^, highlighting the importance of early post-CCU monitoring. However, monitoring every patient discharged from the CCU in the general ward can lead to excessive medical costs and increased burdens on healthcare workers. Thus, identifying patients at high risk of readmission to CCU during their hospital stay is critical. In our previous work, we introduced a risk stratify model to predict readmission to CCU with AUC range 0.70-0.72^2^. Recently, one study using 14 features to develop a recurrent neural network to predict CCU readmission and found the balanced accuracy range from 0.619-0.727^19^. Another studies utilize 41 variables to develop machine learning model and had performance range from 0.744-0.986^20^. It appears that by having more associated features, we are able to build better model on it.

In this study, we developed a machine learning model from an initial set of 40 variables and refined this list to select the best subset of features through stepwise feature selection while training the ML models. The stepwise iteration process was conducted independently for each model, resulting in a tailored set of predictors for each. For instance, while Ventilator use and Length of CCU stay were consistently identified as top features across all models, other variables like Platelet count and Hypertension were selectively included based on the specific model’s algorithmic requirements and performance metrics. Through this approach, we were able to enhance the performance of each model by concentrating on the most informative features, thereby increasing the models’ predictive accuracy and robustness. The resulting subsets of features are expected to provide the basis for more streamlined and targeted predictive tools in a clinical environment, focusing on the most relevant factors for CCU readmission risk.

This stepwise selection approach was preferred over methods like Principal Component Analysis (PCA)^21^ and regression-based selection due to its unique benefits. Unlike PCA, which transforms the feature space into a set of linearly uncorrelated components and might obscure the interpretability of individual features, forward stepwise selection maintains the original features, thus preserving their interpretability^22^. Furthermore, while regression-based techniques might introduce all available features into the model simultaneously, potentially leading to overfitting, the stepwise method selectively incorporates only those features that provide meaningful improvements. The stepwise approach, therefore, achieves a balance between model complexity and predictive power, ensuring the inclusion of features that are both interpretable and relevant to the model’s predictive capacity, an equilibrium that is less readily achieved with PCA or regression-based feature inclusion methods.

The strategic adjustment of the decision threshold significantly impacts our model’s ability to predict CCU readmissions. Setting a threshold of 0.025 enhances our model’s sensitivity without significantly compromising specificity, enabling a more comprehensive detection of at-risk patients, as indicated in Table 3. Conversely, with the threshold at 0.125, our model achieves a high specificity of 0.90 in the hold-out test set, ensuring accurate identification of non-readmissions while accepting a reduced sensitivity of 0.534. A similar pattern of sensitivity and specificity was observed across three other external validation hospitals. These adjustments highlight the delicate balance between capturing true positives and minimizing false alerts, allowing clinicians to tailor the model’s use to the clinical scenario. In the hold-out test set, patients with model predictions greater than 0.125 demonstrated a 7.5-fold increased risk of CCU readmission compared to those with predictions lower than 0.125. In contrast, patients with prediction scores lower than 0.025 had a CCU readmission rate of only 0.5%. The decision on the optimal threshold must be carefully calibrated to the clinical setting, weighing the urgency of detecting readmissions against the consequences of false alarms. A lower threshold, such as 0.025, is prudent in high-stakes environments where a missed readmission could lead to serious consequences, while a higher threshold, like 0.125, may be more suitable in contexts where the costs of false positives are of greater concern.

This study has several limitations. Due to its retrospective nature, data quality issues were present at both the development and external validation sites. Although the external validation hospitals and the primary hospital utilize the same EHR system, which facilitates consistent documentation of included features, assessing the model’s performance in hospitals with different EHR systems poses a challenge. In this study, we excluded patients who died or were transferred to other hospitals, which might introduce survival bias. This could potentially skew the data towards cases with less severity and affect the model’s ability to generalize to sicker patients. Furthermore, the clinical application of this model encounters limitations. The subset of features selected from the GB model primarily consisted of non-modifiable characteristics from a clinician’s perspective. And despite the model’s relatively PPVs, its ability to effectively rule out CCU readmissions remains a valuable asset. It can provide healthcare practitioners with a means to more efficiently allocate resources and enhance the care for patients less likely to experience readmission. Moreover, the implementation of any model into clinical practice necessitates ongoing surveillance and potential recalibration to suit the local context.

## Conclusion

This study demonstrates the potential of ML-driven methods in predicting imminent CCU readmission during the initial hospital stay, leveraging comprehensive datasets from four prominent hospitals. Employing ML models, the research effectively utilized EHR features to optimize prediction performance. The GB model emerged superior, exhibiting remarkable performance with an AUC of 0.879 in the hold-out test set and external validations confirmed the model’s robustness across different patient demographics and settings. Overall, this work reinforces ML’s promise in enhancing patient care, though continuous refinement and validation are essential for clinical implementation.

## Data Availability

The data underlying this article will be shared on reasonable request to the corresponding author.

---

**Supplemental Table 1.**
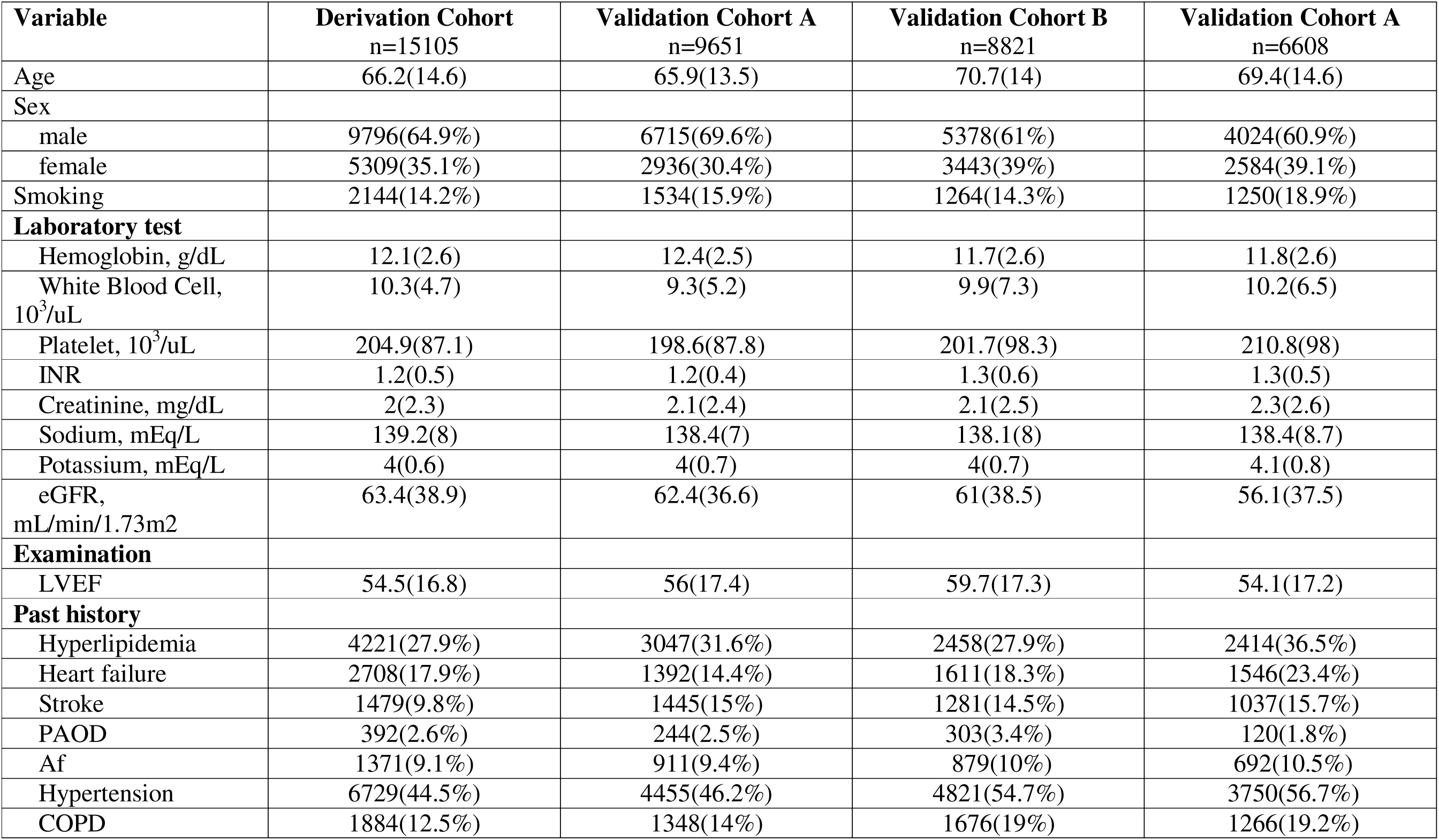
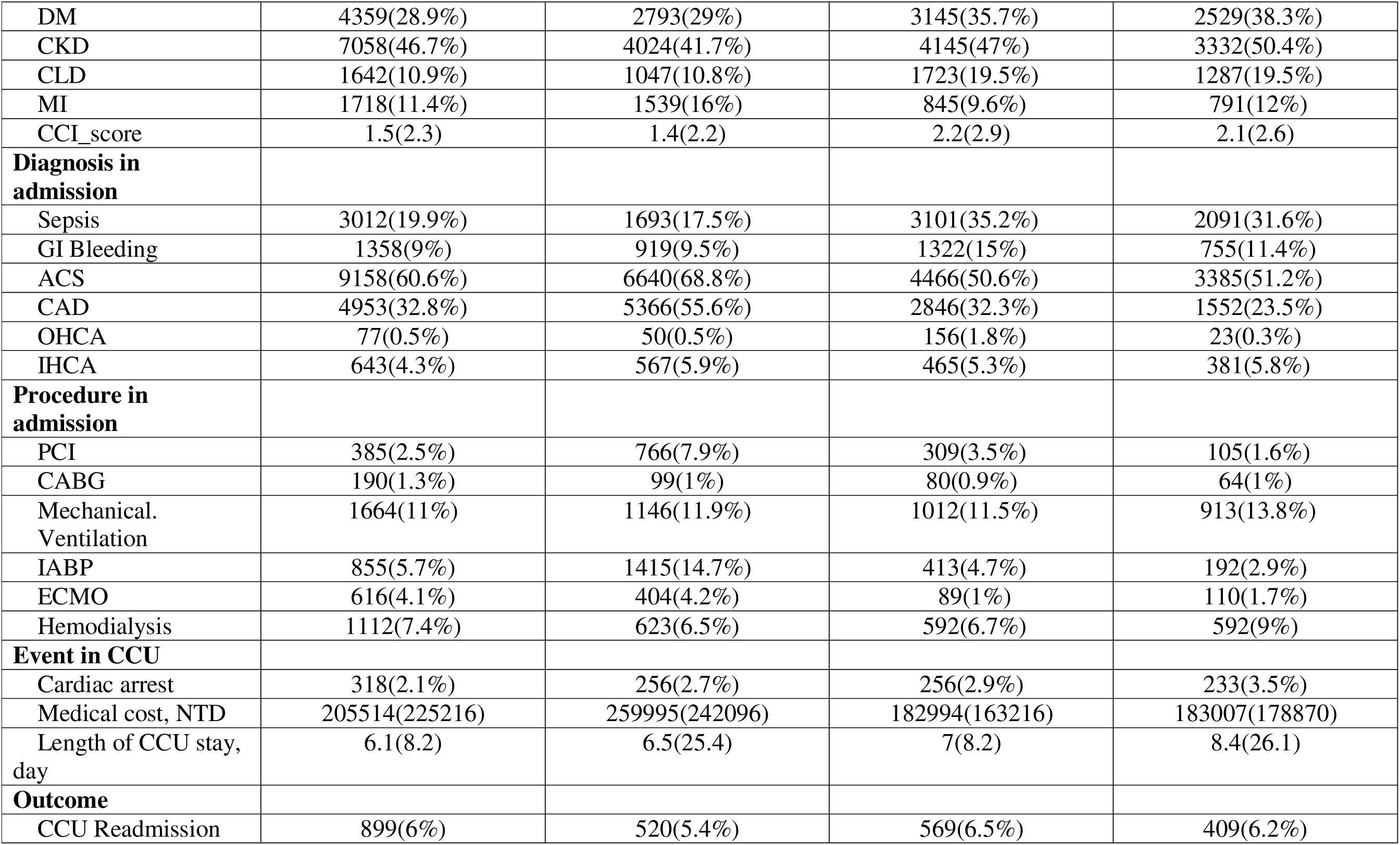
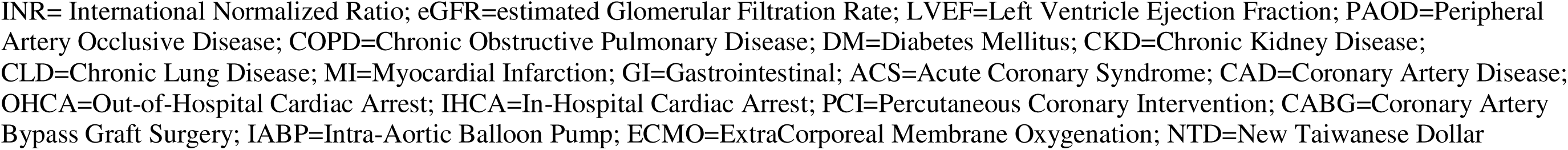
Complete list of variables in selected hospitals.

